# Performance of Rapid Antigen Tests to Detect Symptomatic and Asymptomatic SARS-CoV-2 Infection

**DOI:** 10.1101/2022.08.05.22278466

**Authors:** Apurv Soni, Carly Herbert, Honghuang Lin, Yi Yan, Caitlin Pretz, Pamela Stamegna, Biqi Wang, Taylor Orwig, Colton Wright, Seanan Tarrant, Stephanie Behar, Thejas Suvarna, Summer Schrader, Emma Harman, Chris Nowak, Vik Kheterpal, Lokinendi V Rao, Lisa Cashman, Elizabeth Orvek, Didem Ayturk, Laura Gibson, Adrian Zai, Steven Wong, Peter Lazar, Ziyue Wang, Andreas Filippaios, Bruce Barton, Chad J. Achenbach, Robert L. Murphy, Matthew Robinson, Yukari C. Manabe, Shishir Pandey, Andres Colubri, Laurel O’Connor, Stephenie C. Lemon, Nisha Fahey, Katherine L Luzuriaga, Nathaniel Hafer, Kristian Roth, Toby Lowe, Timothy Stenzel, William Heetderks, John Broach, David D McManus

## Abstract

**Background:** Performance of rapid antigen tests for SARS-CoV-2 (Ag-RDT) varies over the course of an infection, and their performance in screening for SARS-CoV-2 is not well established. We aimed to evaluate performance of Ag-RDT for detection of SARS-CoV-2 for symptomatic and asymptomatic participants.

**Methods:** Participants >2 years old across the United States enrolled in the study between October 2021 and February 2022. Participants completed Ag-RDT and molecular testing (RT-PCR) for SARS-CoV-2 every 48 hours for 15 days. This analysis was limited to participants who were asymptomatic and tested negative on their first day of study participation. Onset of infection was defined as the day of first positive RT-PCR result. Sensitivity of Ag-RDT was measured based on testing once, twice (after 48-hours), and thrice (after 96 hours). Analysis was repeated for different Days Post Index PCR Positivity (DPIPP) and stratified based on symptom-status.

**Results:** In total, 5,609 of 7,361 participants were eligible for this analysis. Among 154 participants who tested positive for SARS-CoV-2, 97 were asymptomatic and 57 had symptoms at infection onset. Serial testing with Ag-RDT twice 48-hours apart resulted in an aggregated sensitivity of 93.4% (95% CI: 89.1-96.1%) among symptomatic participants on DPIPP 0-6. Excluding singleton positives, aggregated sensitivity on DPIPP 0-6 for two-time serial-testing among asymptomatic participants was lower at 62.7% (54.7-70.0%) but improved to 79.0% (71.0-85.3%) with testing three times at 48-hour intervals.

**Discussion:** Performance of Ag-RDT was optimized when asymptomatic participants tested three-times at 48-hour intervals and when symptomatic participants tested two-times separated by 48-hours.

## Introduction

SARS-CoV-2 diagnostic testing remains a cornerstone in our nation’s fight against COVID-19, and at-home rapid antigen tests (Ag-RDTs), while not perfect, provide a fast and convenient testing option. This type of test is available without a prescription (i.e., over-the-counter [OTC]), easy-to-use, widely available, and, in some cases, preferred by the population over molecular assays that require appointments, waiting in-line at testing centers, and waiting 24-48 hours for results.^1–3^ Despite their popularity, key gaps remain in our understanding of these tests, notably their performance as a screening tool among asymptomatic people. Reports on Ag-RDT performance among individuals testing while they are asymptomatic have been highly varied, ranging from sensitivities of 35.8% to 71% in cross-sectional screening evaluations.^4,5^ However, performance has typically been evaluated based on the single use of Ag-RDTs, and few studies have evaluated serial testing performance of Ag-RDTs among asymptomatic individuals.

Furthermore, United States Food and Drug Administration (FDA) emergency use authorization (EUA) of OTC antigen tests required a post-authorization demonstration of Ag-RDT performance in a population with asymptomatic infection using serial testing. This manuscript describes primary findings from a large study designed in coordination with the National Institutes of Health (NIH), Food and Drug Administration (FDA), and three major rapid antigen test manufacturers to evaluate the performance of serial testing using rapid antigen tests for detection of SARS-CoV-2 among asymptomatic individuals within the first week of infection. A primary goal of this study was to provide broadly applicable data that could be leveraged to satisfy the post-authorization requirement for all authorized OTC antigen tests.

## Methods

### Study Population and Design

Between October 18, 2021 and January 31, 2022, this 15-day prospective cohort study enrolled participants over the age of two years from across the country through a novel digital site-less study protocol. Details of the study design and protocol are described elsewhere.^6^ This study was approved by WIRB-Copernicus Group (WCG) Institutional Review Board (20214875). In brief, participants were eligible to enroll through a smartphone app if they did not have a SARS-CoV-2 infection in the prior three months, were without any symptoms in the 14 days prior to enrollment and were able to drop-off prepaid envelopes with nasal swab samples at their local FedEx drop-off location. Enrolled participants were assigned to one of three types of EUA Ag-RDT (Quidel QuickVue At-Home OTC COVID-19 Test, BinaxNow COVID-19 Antigen Self-Test, or BD Veritor At-Home COVID-19 Test) and received a home delivery of 10 Ag-RDTs and 7 home collection kits for reverse transcriptase polymerase chain reaction (RT-PCR) samples. Participants were asked to perform two self-collected bilateral anterior nasal swab collections and paired testing (Ag-RDT (at home) and RT-PCR (mailed to central lab)) between study day 1 and study day 13 on 48 hour intervals, with an additional end-of-study bilateral anterior nasal swab collection for home Ag-RDT on study day Two FDA-authorized, high sensitivity RT-PCR assays were performed on each nasal swab sample received at the central lab, and an additional tiebreaker assay was performed if the two RT-PCR assays were discordant.

### Measures

Ag-RDT results were based on self-reporting (Quidel QuickVue At-Home OTC COVID-19 Test, BinaxNow COVID-19 Antigen Self-Test) or an automatic reader (BD Veritor At-Home COVID-19 Test), as per EUA instructions for use. Molecular comparator RT-PCR results were based on a combination of molecular test results for detection of SARS-Cov-2 infection (Table S1), and onset of infection was defined as the day on which the molecular comparator result was positive for the first time. Cycle threshold (Ct) values for the E-gene from one RT-PCR test were used as a measure to quantify viral load. To approximate performance of Ag-RDT if a person starts testing on different days from onset of infection, we identified Days Past Index PCR Positivity (DPIPP) as different strata, for which, performance was calculated. Symptomatic or asymptomatic classification was based on presence or absence of symptoms on DPIPP for which the performance is calculated. Therefore, an individual who was asymptomatic on DPIPP 0 may become symptomatic on DPIPP 2 and vice-versa.

### Statistical Analysis

Participants were eligible for inclusion in this analysis if they did not report any symptoms and had negative SARS-CoV-2 molecular and Ag-RDT tests on study day 1. Our decision to pool performance across the different tests was based on: 1) findings not shown in this report that suggested the sensitivity of different Ag-RDT was similar to each other as a function of the viral load; and 2) the study was not designed to evaluate differences in performance between the three different types of Ag-RDTs. Performance was calculated using sensitivity (rapid antigen positivity/comparator positivity) for single-day testing, two-times serial testing at 48-hour intervals, and three-times serial testing at 48-hour intervals for symptomatic and asymptomatic individuals based on day and patterns of positivity, as described in Table 1. Calculations for sensitivity were repeated with testing starting on different DPIPP. Confidence intervals for one-week sensitivity were estimated using the bootstrapping technique.^7^ Negative Percent Agreement was calculated as a proportion of paired tests (Ag-RDT and molecular tests performed on the same-day) based on the following formula: True Negative/[False Positive + True Negative], where True Negative refers to instances a negative molecular test was paired with a negative Ag-RDT on the same day and False-Positive refers to a negative molecular test paired with a positive Ag-RDT test. To evaluate the viral dynamics during infection, a mixed effects logistic regression model was used to predict the probability of Ag-RDT positivity based on Ct value and symptom status. Ct values, symptom status, rapid antigen testing series (one test, two-tests, or three-tests), and their three-way interactions were fixed effects, while participants’ IDs was included as a random effect. Additionally, we used the Cochran-Armitage test for trend to evaluate the associations between symptoms and Ct value categories (i.e., <20, 20-24.99, 25-29.99, 30-34.99, or 35+) among individuals who tested positive on a single rapid antigen test, two consecutive tests, and three consecutive tests, respectively. We used Bonferroni corrections (0.05/3=0.017) to adjust for multiple comparisons. All analyses were performed using R 4.1.1.^8^

**Table 1:** Matrix for Calculation of Sensitivity of Rapid Antigen Tests for Detection of SARS-CoV-2 Virus in Relation to Days Past Index PCR Positivity (DPIPP)

| Days Past Index PCR Positivity – DPIPP (strata) | Denominator for Sensitivity: PCR Positive on following DPIPP | Numerator for Sensitivity: At least one positive rapid antigen test on one of the following DPIPP<br>(Other days could not be missing, invalid, or uninterpreted) |  |  |
| --- | --- | --- | --- | --- |
|  |  | Single test | 2-test serial testing | 3-test serial testing |
| 0 | 0 | 0 | 0, 2 | 0, 2, 4 |
| 2 | 0 and 2 | 2 | 2, 4 | 2, 4, 6 |
| 4 | 0 and 4 | 4 | 4, 6 | 4, 6, 8 |
| 6 | 0 and 6 | 6 | 6, 8 | 6, 8, 10 |
| 8 | 0 and 8 | 8 | 8, 10 | 8, 10, 12 |
| 10 | 0 and 10 | 10 | 10, 12 | NA |
| Classification of symptomatic vs asymptomatic based on symptom self-report on DPIPP |  |  |  |  |

## Results

A total of 7,361 participants enrolled in the study, and 5,353 were eligible for this analysis. Of the participants eligible for analysis, 154 individuals tested RT-PCR positive for SARS-CoV-2 during the study based on a composite definition described in Table S1; 97 were without symptoms and 57 had symptoms at infection onset (Figure 1). Among the 5,199 participants who did not test positive, there were 32,998 days of paired Ag-RDT and RT-PCR testing, where comparator result was negative. Among these, 32,862 days had a concordant Ag-RDT negative result, yielding a negative percent agreement of 99.6% (95% CI: 99.5-99.7%).

**Figure 1:**
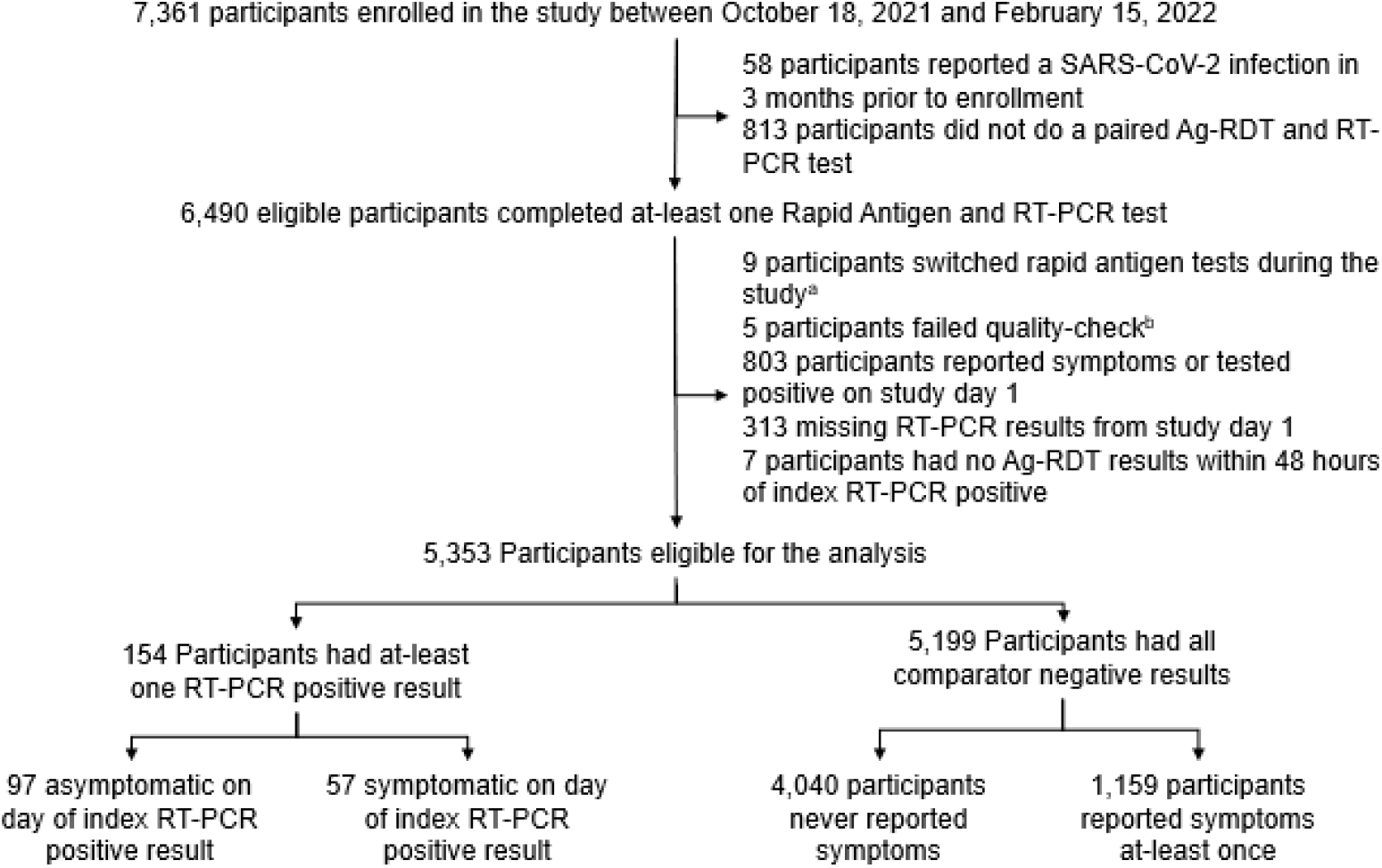
CONSORT Diagram for Test Us At Home Study to Calculate Performance of Rapid Antigen Tests for Detection of SARS-CoV-2 Virus. 7,361 total participants were enrolled in the study, and 154 participants were eligible for the analysis and tested positive for SARS-CoV-2 by RT-PCR during the study period. Of these participants, 97 were asymptomatic and 57 were symptomatic on day of index comparator positive result. a= participants replaced their assigned rapid antigen tests with commercially obtained rapid antigen tests; b= dates of RT-PCR testing could not be verified based on triangulation of self-reported, shipping, and resulting data.

Performance of Ag-RDT to detect SARS-CoV-2 on day of infection onset (DPIPP: 0) was higher among symptomatic participants (59.6%, 46.7-71.4%) in comparison to asymptomatic participants (9.3%, 5.0-16.7%) (Figure 2; Table S2). Serial-testing with two Ag-RDT tests 48-hours apart (Symptomatic: 92.2%, 81.5-96.9%; Asymptomatic: 39.3%, 29.8-49.7%) and three Ag-RDT tests 48-hours apart (Symptomatic: 93.6%, 82.8-97.8%; Asymptomatic: 56.4%, 45.4-66.9%) improved performance of Ag-RDT.

**Figure 2:**
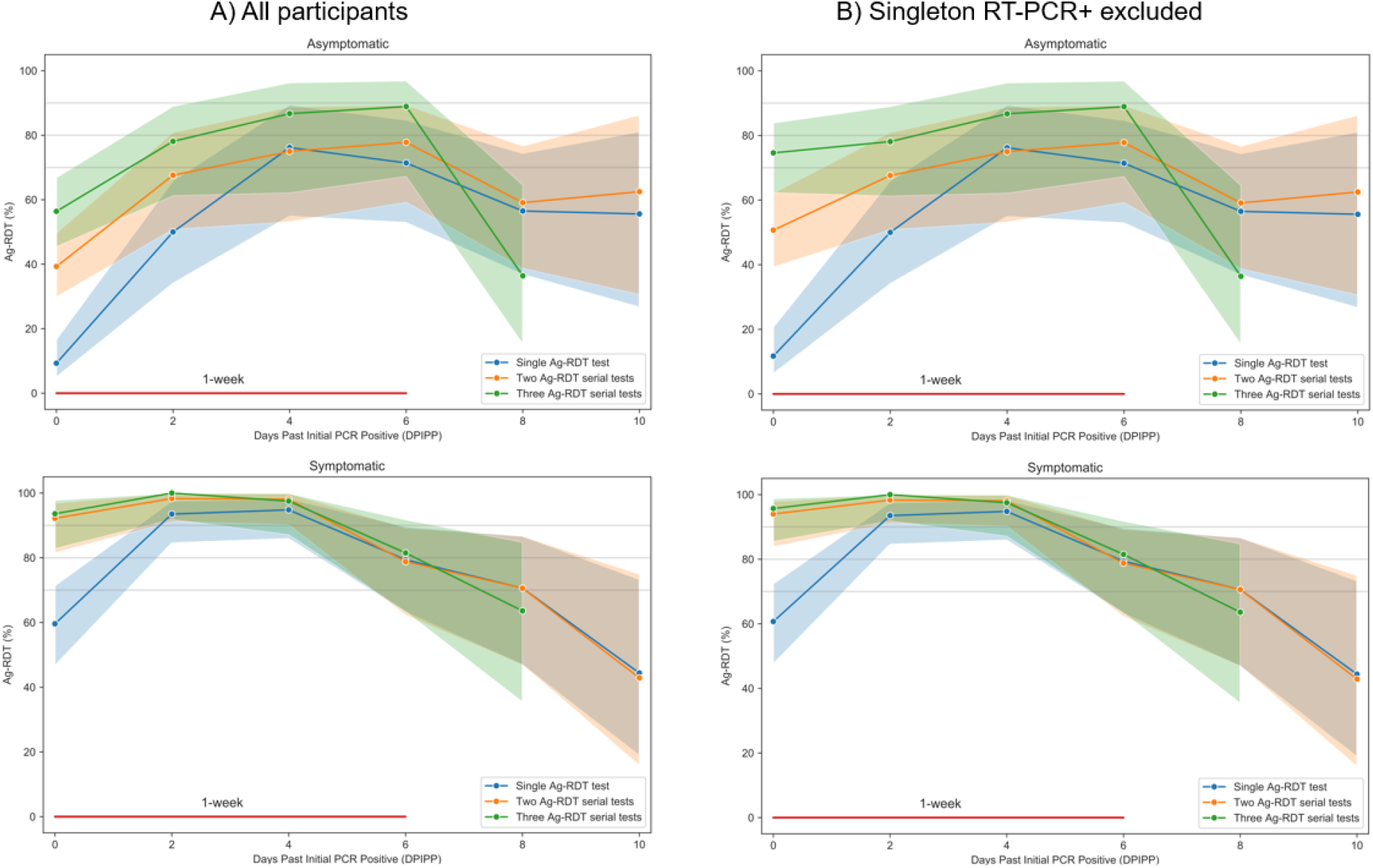
Performance of Rapid Antigen Tests for Detection of SARS-CoV-2 Virus in Relation to First Day of Molecular Positivity. Performance was calculated using sensitivity (rapid antigen positivity/comparator positivity) for single-day testing, two-times serial testing at 48-hour intervals, and three-times serial testing at 48-hour intervals for symptomatic and asymptomatic individuals based on day and patterns of positivity. Calculations for sensitivity were repeated with testing starting on different DPIPP. Error bars represent 95% Confidence Intervals. Performance of Ag-RDT to detect SARS-CoV-2 on day of infection onset was higher among symptomatic participants (59.6%) compared to asymptomatic participants (9.3%). Serial-testing with two Ag-RDT tests 48-hours apart and three Ag-RDT tests 48-hours apart improved performance of Ag-RDT within the first week of SARS-CoV-2 infection. Excluding participants with singleton RT-PCR+ results improved sensitivity of Ag-RDT among asymptomatic participants to 11.7%, 50.7%, and 74.6% based on testing one, two, or three times with Ag-RDT at 48-hour intervals, respectively, beginning on DPIPP 0.

Notably, we observed that twenty participants had a singleton RT-PCR+, defined as a positive test preceded and followed by a negative RT-PCR test within 48-54 hours. Of those with singleton RT-PCR+ tests, none tested positive on Ag-RDT, only one had symptoms on the day of the singleton positive test, and average Ct value was >35 (Figure S1). Excluding these participants did not impact sensitivity of Ag-RDT among symptomatic participants, but improved asymptomatic sensitivity to 11.7%, 50.7%, and 74.6% based on testing one, two, or three times with Ag-RDT at 48-hour intervals, respectively, beginning on DPIPP 0.

To approximate real-world scenarios, where a person may not necessarily start testing with Ag-RDT on the day of infection onset, we calculated performance separately on DPIPP 2, 4, 6, 8, and 10 (Figure 2; Table S2) to approximate scenarios where a person started serially testing with Ag-RDTs on those days. Aggregated performance of Ag-RDT for DPIPP 0-6 among all participants who were symptomatic on a given DPIPP was 82.5% (78.3-86.3%) for single-timepoint testing but with a range of sensitivity (calculated on a per DPIPP day basis) from 59.6 to 94.8%. Serial-testing using two-time testing improved sensitivity to 93.4% (90.4-95.9%) and 94.3% (91.4-97.0%) for three-time testing. Sensitivity of performing single test, two-test serial testing, and three-test serial testing for asymptomatic people was 34.4% (28.8-39.8%), 55.3% (48.2-61.6%), 68.5% (61.0-75.7%), respectively during the first week of infection (DPIPP 0-6). Excluding singleton RT-PCR positive, the first-week (DPIPP 0-6) sensitivity for asymptomatic individuals was 38.8% (32.7-45.2-%), 62.7% (57.0-70.5%), 79.0% (70.1-87.4%), respectively for testing one, two, or three times with Ag-RDT at 48-hour intervals.

Performance of Ag-RDT among symptomatic and asymptomatic participants was evaluated by Ct value to analyze the performance by viral load. The distribution of Ct values significantly differed between symptomatic and asymptomatic participants, with symptomatic participants having lower Ct values on average than asymptomatic participants at DPIPP 0 and 2 (p<0.001) (Figure 3). On the day of index PCR positivity, more than 75% of asymptomatic individuals had a Ct value 30 or higher, whereas less than 33% of symptomatic individuals had a Ct value 30 or higher. At the end of 1-week from index PCR positivity (DPIPP 6), the majority of both asymptomatic and symptomatic individuals had a Ct value less than 30. Figure 4 demonstrates the sensitivity of Ag-RDT for different Ct values based on postestimation results from a multilevel model. The sensitivity was lowest among asymptomatic participants who performed a single test for all Ct values > 20, compared to symptomatic participants who performed a single test and symptomatic and asymptomatic participants who performed two-test and three-test serial testing (Figure 4; Table S3). Two-test serial testing among symptomatic individuals and three-times serial testing among both asymptomatic individuals and symptomatic individuals demonstrated sensitivity above 80% for Ct values < 32.

**Figure 3:**
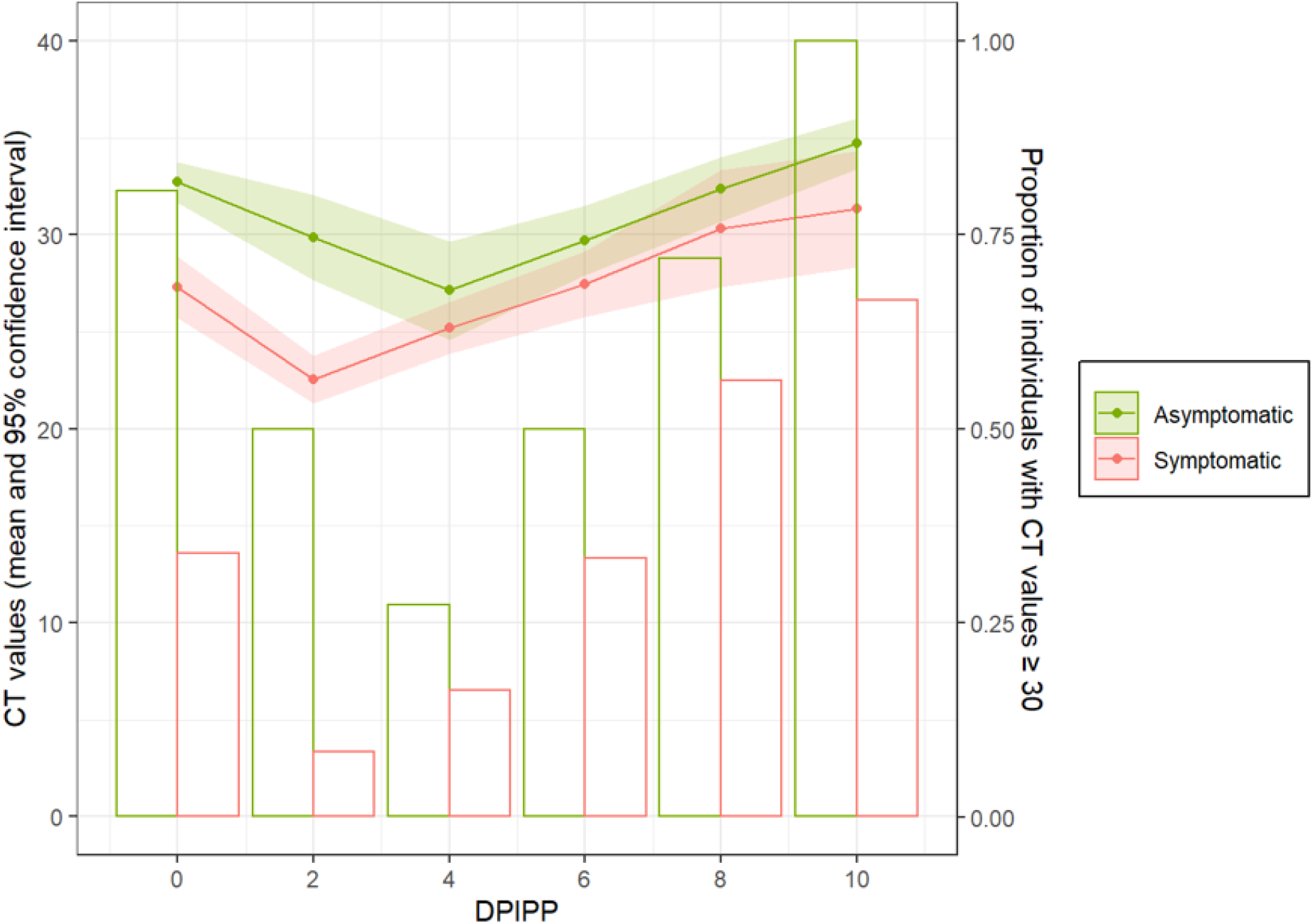
Cycle Threshold values by Day Post Index PCR-Positivity by Symptom Status. Error bars represent 95% Confidence Intervals. Symptomatic participants who tested positive by RT-PCR had significantly lower Cycle threshold (Ct) values on average than asymptomatic participants at DPIPP 0 and 2. On DPIPP 0, >75% of asymptomatic individuals had a Ct value ≥30, whereas <33% of symptomatic individuals had a Ct value ≥30. Symptomatic participants had a lower proportion of individuals with CT values ≥30 at all DPIPP. DPIPP: Day post index PCR-positive.

**Figure 4:**
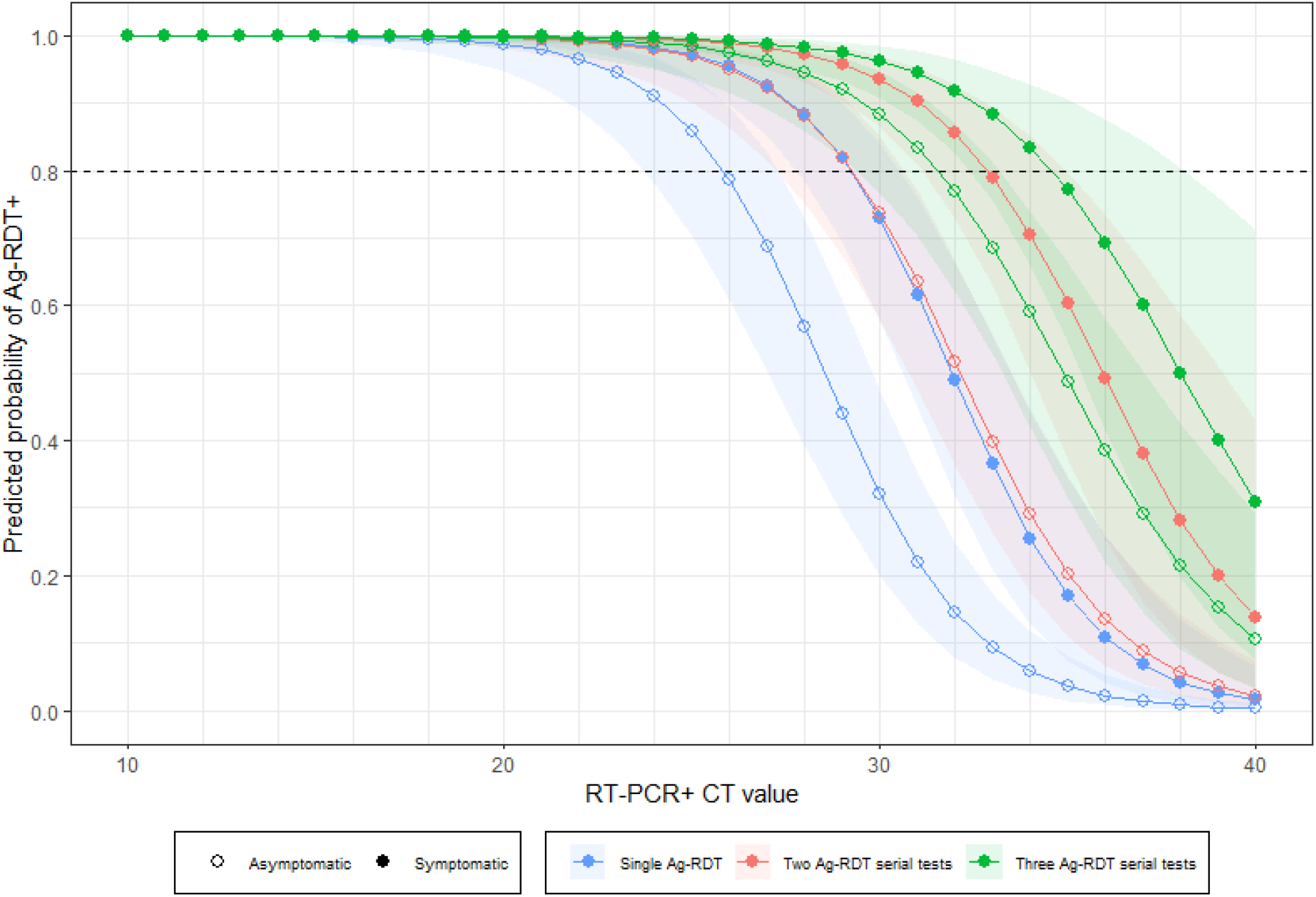
Predicted Probability of Rapid Antigen Test Positivity by Symptom Status and Serial Testing Schedule. A mixed effects logistic regression model was used to predict the probability of Ag-RDT positivity based on Ct value and symptom status. Error bars represent 95% Confidence Intervals. The sensitivity was lowest among asymptomatic participants who performed a single test for all Ct values > 20. Two-test serial testing among symptomatic individuals and three-times serial testing among both asymptomatic individuals and symptomatic individuals demonstrated sensitivity above 80% for Ct values < 32.

## Discussion

We report findings from the largest study to date of paired Ag-RDT and RT-PCR testing for a comparative performance evaluation of Ag-RDTs among people with and without symptoms. These results provide compelling reasons to suggest that frequency of Ag-RDT testing should be adjusted to include additional repeat testing. These data suggest an improvement in test performance when symptomatic individuals test two times 48-hours apart using Ag-RDTs. Likewise, there are notable performance improvements in asymptomatic individuals when an initial Ag-RDT test was followed by at least 2 subsequent tests at 48-hour intervals. These results should be considered in the context of our study protocol which indicated testing at 48-hour intervals, and thus these data cannot support conclusions about serial testing for time intervals shorter than 48-hours.

These findings represent a comprehensive evaluation of the time dependent performance of Ag-RDT tests among the intended use population (i.e., symptomatic and asymptomatic individuals) throughout the course of molecular test positivity. Restricting findings from our study to match observation windows from previous studies, we found similar sensitivity as found in previous studies for asymptomatic and symptomatic participants for a single-time test.^9^ Unlike previous reports, which used composite sampling methods and lacked sufficient longitudinal data to adequately evaluate performance of Ag-RDT from the onset of infection, we were able to approximate performance of Ag-RDT for symptomatic and asymptomatic users by comparing performance within the first week of infection, to align with the indications listed in the EUA, and to evaluate the performance of serial testing within this paradigm.^10,11^

We observed that more than one in ten new infections were singleton-positive, which escaped detection by rapid antigen tests. Evaluation of Ct values of these singleton-positive revealed that all of them had Ct value > 30 and average Ct count was 35. The finding of singleton RT-PCR positive testing needs to be further investigated to understand the clinical significance of this observation.

Our finding of higher Ct value associated with lower sensitivity is in line with results from a comprehensive meta-analysis encompassing data from 214 clinical studies and 112,323 samples, which demonstrated that sensitivity of rapid antigen testing deteriorated with increasing Ct values.^9^ We also observed that rapid antigen tests have higher sensitivity among symptomatic participants, regardless of Ct value. The finding that performance of rapid antigen tests differed with respect to Ct values between symptomatic and asymptomatic participants was unexpected, as rapid antigen test performance has often been considered to be a function of viral load.^12,13^ However, this may suggest that the difference in performance between asymptomatic and symptomatic participants is more than just a function of viral dynamics. It is possible that symptomatic and asymptomatic individuals differ in three other domains: 1) interpretation of results; 2) administration of tests; 3) physiology (i.e., amount of secretions available for sampling). Previous work found that symptom status was not a predictor of false negative results; however, self-interpretation of results may introduce bias.^14^ These three hypotheses are subject to further inquiry, as it is important to determine the role of these factors in rapid antigen test performance. Additionally, a previous report suggested that certain haplotypes of the human leukocyte antigen (HLA) loci are two to eight-fold more likely to experience an asymptomatic infection, and in a large GWAS study, this haplotype was found to be prevalent in roughly 10% of the patients.^15^ Similar observations have been made with Human Immunodeficiency Virus, where certain genotypes are associated with lower propensity of infection. The impact of HLA haplotype on COVID-19 infections should be investigated further. This study does have limitations. This study was conducted during the circulation of the Delta and Omicron SARS-CoV-2 variants, and future variants may warrant further investigation, especially as milder, less symptomatic variants emerge.^16,17^ Additionally, specimens were self-collected for RT-PCR and Ag-RDTs were self-performed. However, data has consistently shown substantial agreement between self-collected and clinician-collected anterior nasal swabs for SARS-CoV-2 testing.^18,19^ Further, this primary analysis does not account for differences in severity or type of symptoms. Public health implications of our findings suggest that people testing for SARS-CoV-2 should exercise caution despite an initial negative rapid antigen-test and favor mask-wearing and avoiding crowded places if they suspect they may be infected or have been exposed. Additionally, the rates of false positive results in the study were low; therefore, any Ag-RDT positive result should be considered positive without the need to retest. Further, in the context of reports of viral culture positivity more than five days after initial positive test, our findings support isolation for a longer period of time to prevent the potential of spread of SARS-CoV-2 to others.^20^ Further research is needed to quantitatively estimate the benefits of Ag-RDT for early detection of infection and initiation of treatment, especially in settings where access to molecular testing is limited or molecular test results are delayed. Dissemination of clear guidance for appropriate testing using Ag-RDT based on data from this study may help preserve confidence in the performance of serial Ag-RDT to detect SARS-CoV-2 virus, especially as reports of individual false negative Ag-RDT from inadequate serial testing, contrary to the tests’ intended usage and guidance from the FDA, proliferate in lay media.

## Data Availability

All data produced are available online at https://github.com/soni-lab.

## Acknowledgment

We are grateful to our study participants and to our collaborators from the National Institute of Health (NIBIB and NHLBI) who provided scientific input into the design of this study and interpretation of our results but could not formally join as co-authors due to institutional policies and to the Food and Drug Administration (Office of In Vitro Diagnostics and Radiological Health) for their involvement in the primary TUAH study. We received meaningful contributions from Drs. Bruce Tromberg, Jill Heemskerk, Dennis Buxton, Felicia Qashu, Erin Iturriaga, Jue Chen, Andrew Weitz, and Krishna Juluru. We are greatly appreciative of the contribution to this study by the numerous staff at UMass Chan, including critical support from Karen Gilliam, Mary Janet McCarthy, Amber Showers, Cynthia Kinahan, Kimberly Cantin, and Danielle Howard. We would also like to acknowledge the support provided by clinical coordinators from Threewire, Inc. We are thankful to county health departments across the country who helped with recruitment for this study by spreading the word in their networks.

