## Supplemental Appendix for "Performance of Rapid Antigen Tests to Detect Symptomatic and Asymptomatic SARS-CoV-2 Infection"

### Supplemental Tables:

Supplementary Table 1: Definition of Positive Comparator Test Based on Molecular Assays

| Roche cobas SARS-CoV-2 Test | Quest SARS-CoV-2 RT-PCR | Hologic Aptima SARS-CoV-2 Assay | Comparator |
|---|---|---|---|
| DETECTED | DETECTED | Other | Positive |
| DETECTED | Other | DETECTED | Positive |
| Other | DETECTED | DETECTED | Positive |
| DETECTED | DETECTED | DETECTED | Positive |

Other: NOT DETECTED, INVALID, or ASSAY NOT PERFORMED
If Roche and Quest LDT were discordant and Aptima result was missing or not performed due to insufficient sample, that data point was excluded from the analysis

Supplementary Table 2: Sensitivity of Rapid Antigen Tests (Ag-RDT) based on symptom status on different Days Past Index PCR Positivity (DPIPP)

| TIME | SYMPTOMATIC (ALL PARTICIPANTS) | | | ASYMPTOMATIC (ALL PARTICIPANTS) | | | SYMPTOMATIC (EXCLUDES SINGLETON PCR+) | | | ASYMPTOMATIC (EXCLUDES SINGLETON PCR+) | | |
|---|---|---|---|---|---|---|---|---|---|---|---|---|
| | 1x test | 2x test | 3x test | 1x test | 2x test | 3x test | 1x test | 2x test | 3x test | 1x test | 2x test | 3x test |
| DPIPP 0 | 34/57 59.6 (46.7,71.4) | 47/51 92.2 (81.5,96.9) | 44/47 93.6 (82.8,97.8) | 9/97 9.3 (5.0,16.7) | 35/89 39.3 (29.8,49.7) | 44/78 56.4 (45.4,66.9) | 34/56 60.7 (47.6,72.4) | 47/50 94.0 (83.8,97.9) | 44/46 95.7 (85.5,98.8) | 9/77 11.7 (6.3,20.7) | 35/69 50.7 (39.2,62.2) | 44/59 74.6 (62.2,83.9) |
| DPIPP 2 | 58/62 93.5 (84.6,97.5) | 59/60 98.3 (91.1,99.9) | 43/43 100.0 (91.8,100.0 | 17/34 50.0 (34.1,65.9) | 23/34 67.6 (50.8,80.9) | 25/32 78.1 (61.2,89.0) | 58/62 93.5 (84.6,97.5) | 59/60 98.3 (91.1,99.9) | 43/43 100.0 (91.8,100.0) | 17/34 50.0 (34.1,65.9) | 23/34 67.6 (50.8,80.9) | 25/32 78.1 (61.2,89.0) |
| DPIPP 4 | 55/58 94.8 (85.9,98.2) | 53/54 98.1 (90.2,99.9) | 39/40 97.5 (87.1,99.9) | 16/21 76.2 (54.9,89.4) | 15/20 75.0 (53.1,88.8) | 13/15 86.7 (62.1,96.3) | 55/58 94.8 (85.9,98.2) | 53/54 98.1 (90.2,99.9) | 39/40 97.5 (87.1,99.9) | 16/21 76.2 (54.9,89.4) | 15/20 75.0 (53.1,88.8) | 13/15 86.7 (62.1,96.3) |
| DPIPP 6 | 27/34 79.4 (63.2,89.7) | 26/33 78.8 (62.2,89.3) | 22/27 81.5 (63.3,91.8) | 20/28 71.4 (52.9,84.7) | 21/27 77.8 (59.2,89.4) | 16/18 88.9 (67.2,96.9) | 27/34 79.4 (63.2,89.7) | 26/33 78.8 (62.2,89.3) | 22/27 81.5 (63.3,91.8) | 20/28 71.4 (52.9,84.7) | 21/27 77.8 (59.2,89.4) | 16/18 88.9 (67.2,96.9) |
| DPIPP 8 | 12/17 70.6 (46.9,86.7) | 12/17 70.6 (46.9,86.7) | 7/11 63.6 (35.4,84.8) | 13/23 56.5 (36.8,74.4) | 13/22 59.1 (38.7,76.7) | 4/11 36.4 (15.2,64.6) | 12/17 70.6 (46.9,86.7) | 12/17 70.6 (46.9,86.7) | 7/11 63.6 (35.4,84.8) | 13/23 56.5 (36.8,74.4) | 13/22 59.1 (38.7,76.7) | 4/11 36.4 (15.2,64.6) |
| DPIPP 10 | 4/9 44.4 (18.9,73.3) | 3/7 42.9 (15.8,75.0) | | 5/9 55.6 (26.7,81.1) | 5/8 62.5 (30.6,86.3) | | 4/9 44.4 (18.9,73.3) | 3/7 42.9 (15.8,75.0) | | 5/9 55.6 (26.7,81.1) | 5/8 62.5 (30.6,86.3) | |
| DPIPP 0-6 | 174/211 82.5 (78.3,86.3) | 185/198 93.4 (90.4,95.9) | 148/157 94.3 (91.4,97.0) | 62/180 34.4 (28.8,39.8) | 94/170 55.3 (48.2,61.6) | 98/143 68.5 (61.0,75.7) | 174/210 82.9 (78.3,87.9) | 185/197 93.9 (90.7,96.8) | 148/156 94.9 (91.4,97.7) | 62/160 38.8 (32.7,45.2) | 94/150 62.7 (57.0,70.5) | 98/124 79.0 (70.1,87.4) |
| DPIPP 8-10 | 16/26 61.5 (42.5,77.6) | 15/24 62.5 (42.7,78.8) | 22/27 81.5 (63.3,91.8) | 18/32 56.2 (39.3,71.8) | 18/30 60.0 (42.3,75.4) | 16/18 88.9 (67.2,96.9) | 16/26 61.5 (42.5,77.6) | 15/24 62.5 (42.7,78.8) | 7/11 63.6 (35.4,84.8) | 18/32 56.2 (39.3,71.8) | 18/30 60.0 (42.3,75.4) | 4/11 36.4 (15.2,64.6) |

Supplementary Table 3: Sensitivity of Rapid Antigen Tests (Ag-RDT) based on symptom status and Cycle threshold values

| **Ct values** | **Symptomatic** | | | **Asymptomatic** | | |
|---|---|---|---|---|---|---|
| | **1x test** | **2x test** | **3x test** | **1x test** | **2x test** | **3x test** |
| **<20** | 38/38 100% (90.8%-100%) | 38/38 100% (90.8%-100%) | 38/38 100% (90.8%-100%) | 5/5 100% (56.6%-100%) | 6/6 100% (61%-100%) | 6/6 100% (61%-100%) |
| **20-24.99** | 70/71 98.6% (92.4%-99.9%) | 71/71 100% (94.9%-100%) | 71/71 100% (94.9%-100%) | 23/26 88.5% (71%-96%) | 24/26 92.3% (75.9%-97.9%) | 25/26 96.2% (81.1%-99.8%) |
| **25-29.99** | 47/60 78.3% (66.4%-86.9%) | 54/59 91.5% (81.6%-96.3%) | 52/55 94.5% (85.1%-98.1%) | 27/38 71.1% (55.2%-83%) | 33/37 89.2% (75.3%-95.7%) | 33/35 94.3% (81.4%-98.4%) |
| **30-34.99** | 20/41 48.8% (34.3%-63.5%) | 33/41 80.5% (66%-89.8%) | 31/37 83.8% (68.9%-92.3%) | 17/67 25.4% (16.5%-36.9%) | 34/64 53.1% (41.1%-64.8%) | 35/52 67.3% (53.8%-78.5%) |
| **35+** | 6/17 35.3% (17.3%-58.7%) | 8/16 50% (28%-72%) | 10/14 71.4% (45.4%-88.3%) | 5/64 7.8% (3.4%-17%) | 14/57 24.6% (15.2%-37.1%) | 23/49 46.9% (33.7%-60.6%) |

### Supplemental Figures:

Supplemental Figure 1: Cycle Threshold Values of Singleton RT-PCR positive tests

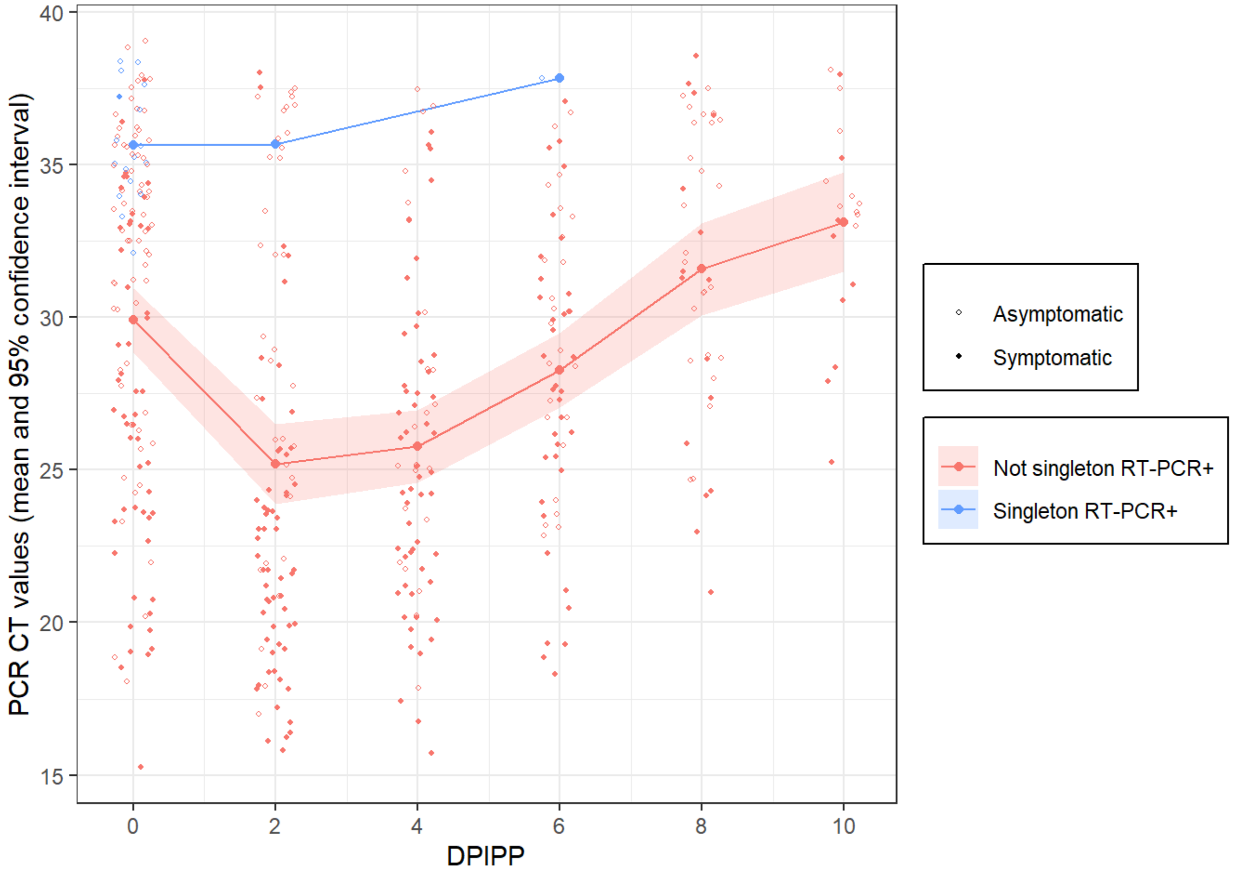
